# Evaluation and Application of a Probability of Success (POS) Framework in Oncology Trials

**DOI:** 10.1101/2025.07.03.25330814

**Authors:** Alex Ziyu Jiang, Yulia Sidi, Xiang Peng, Thomas Jemielita, Sabrina Wan

## Abstract

Oncology drug development encounters considerable challenges due to positive Phase I trials rarely leading to regulatory approvals, extremely competitive landscape, and high financial burdens associated with research and development. To enhance decision-making in this area and address high costs and risks associated with late-stage oncology trials, we developed a Bayesian decision-making framework by enhancing existing mWethodologies for estimating the probability of success (PoS) of these trials. We proposed a hierarchical Bayesian model that incorporates mixture prior weights from estimated benchmark probability of approval using machine learning methods, observed treatment effect from earlier studies, and the design features of Phase III trials for PoS estimation. We illustrated the adaptability of our framework to scenarios where prior data is available only from a single-arm study or when the earlier study pertain to endpoints that differ from the primary endpoint intended for the pivotal trial. We discovered through case and simulation studies that multiple factors affect the PoS predictions, including treatment effect uncertainty from earlier studies; and assessed the robustness of our model by varying the assumed correlation between the early study endpoint and the pivotal study’s primary endpoint is important. Our framework was designed to facilitate a consistent evaluation of proposed oncology clinical trial designs regarding their statistical success, either within a specific program or throughout a pipeline portfolio. We anticipate that this framework will help streamline decision-making process for oncology pivotal trials from trial sponsor’s side, improve drug development efficiency and ultimately benefit patients.

## 1 Introduction

Oncology drug development presents significant challenges with only 3.4% to 6.7% of Phase I trials ultimately resulting in regulatory approvals (Wong et al., 2019; Hay et al., 2014). This rate is notably lower than the recently reported approval likelihood of 14.31% across all therapeutic areas (Schuhmacher et al., 2025). The unique nature of oncology indications which often involve high unmet medical needs and occasionally rare diseases, along with the diverse therapeutic mechanisms, lead to a highly competitive drug development landscape that differs substantially from non-oncology fields. For example, while in many therapeutics areas two successful pivotal trials are required in order to proceed with a regulatory submission, in oncology one pivotal trial is usually sufficient (Food and Drug Administration (FDA), 2021). In addition, in oncology it is common to initiate a pivotal trial based on early endpoint such as objective response rate (ORR) or pathological complete response (pCR) from Phase I or Phase I/II single arm studies, if substantial evidence of anti-tumor activity is observed and the safety profile is manageable. This adds onto the uncertainties in evaluating efficacy in a pivotal study as assessed by long-term endpoint such as overall survival (OS). In addition, the financial burden of oncology research and development (R&D) is significant, with reported expenditures often the highest among various therapeutic areas (Schlander et al., 2021). The escalating costs, highly competitive environment, and frequently limited evidence available at the outset of pivotal trials underscores the necessity for a quantitative approach that integrates prior evidence with the pivotal study design to predict trial success.

Over the past two decades, various approaches addressing the probability of success (PoS) topic in drug development have emerged Wang et al., 2013; Saint-Hilary et al., 2018. Recently, Fougeray et al. (2024) constructed an informative prior for an early futility analysis of the long-term primary endpoint by exploiting the relationship between the primary endpoint and a surrogate endpoint; Proper et al. (2024) proposed a modeling framework based on the propensity-score-based meta-analytic predictive (PS-MAP) prior for the Phase II control arm, in order to objectively incorporate the information at the patient level. Also, Chen et al. (2023) expanded the concept of PoS to pre-clinical and early clinical development, where they proposed the probability of pharmacological success (PoPS) as a tool to inform evidence-based rational progression decisions for early-stage drug candidates by integrating multi-dimensional data. Collectively, these advancements illustrate the evolving landscape of methodologies aimed at enhancing decision-making in drug development.

In this paper, we build upon the previous work by (Hampson et al., 2022) to provide a framework for predicting the success of oncology pivotal trials and assessed its statistical property by varying model parameters. We define trial success as achieving statistical significance for the primary endpoint. Although statistical significance may not solely determine regulatory approval, the success rate for approvals following a new drug application (NDA) or biologics license application (BLA) in oncology (usually after achieving statistical significance in pivotal trials) is reported to be 92% (IQVIA report, 2024). Our approach mirrors that of (Hampson et al., 2022), starting with industry benchmarks to generate an initial estimate of success rate given general characteristics of the pivotal study, such as specific indication and line of therapy using a statistical learning model (Step 1). Step 2 then synthesizes the benchmark PoS, any available Phase I/II data that led to the discussion of a subsequent pivotal trial, and the pivotal trial design through a Bayesian hierarchical model to estimate the proposed trial’s PoS. This step enhances the initial estimate from Step 1 by integrating the observed treatment effects gathered thus far along with uncertainty associated with it. Additionally, it takes into account the specific parameters of the proposed Phase III study design, such as the number of target events, the planned number of interim analyses, and other relevant factors. By integrating this information, Step 2 facilitates a more nuanced revision of PoS, ensuring it is better aligned with real-world data and the unique characteristics of the upcoming study. We emphasize that the primary objective of pivotal study PoS is to enable systematic comparisons among various compounds and studies, thereby equipping decision-makers with a tool to make more informed choices based on the available information.

The remainder of the paper is organized as follows: In Section 2, we outline our approach for estimating PoS. In Section 3, we provide additional considerations for PoS estimation. In Section 4, we present a case study based on past study designs. Finally, we discuss the potential further developments and limitations of our proposal.

## 2 Methods

We assume that *θ*_*P* 3_ is a treatment effect on a primary endpoint *P* in a Phase III study. For example, if *P* denotes the progression-free survival (PFS), *θ*_*P* 3_ is the log hazard ratio (HR) of PFS. The subscript 3 in this notation denotes the phase of the study (i.e., Phase III). In addition, we assume that treatment effect on either the same endpoint *P* or a different endpoint *D* is observed in an earlier Phase I/II or Phase II study which preceded the pivotal trial, defined as *θ*_*P* 2_ and *θ*_*D*2_ respectively . The study level model for these parameters are introduced next.

### 2.1 Study Level Model

The study level model for an observed treatment effect on endpoint *P* in the *k*-th earlier study (*k* = 1, … , *K*, where *K* is the total number of prior studies) is denoted as 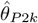 and is assumed to follow a Normal distribution:

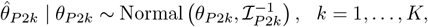

where *θ*_*P* 2*k*_ represents the mean treatment effect on *P* in the *k*-th earlier study, and ℐ_*P* 2*k*_ represents the Fisher information for *θ*_*P* 2*k*_. We further assume a Normal distribution prior for *θ*_*P* 2_:

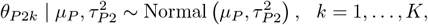

where *µ*_*P*_ is the population level parameter for the treatment effect on *P* . *τ*_*P* 2_ is a hyper-parameter that characterizes the degree of heterogeneity in the treatment effect on *P* across different earlier studies. We assume that *τ*_*P* 2_ is a weakly informative prior with a half-Normal distribution, i.e., 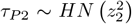 following similar approach to Hampson et al. (2022). Similar to the above, we let *θ*_*P* 3*k*_ be the treatment effect on *P* at the Phase III study and assume it follows a Normal distribution:

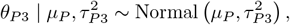

where *τ*_*P* 3_ is, again, assumed to follow a half-Normal distribution, i.e., 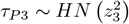. For the purpose of this paper, we assume that 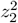 and 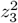 represent “small” and a “very small” heterogeneity in the mean treatment effect across earlier studies and Phase III study, respectively (Hampson et al., 2022). These assumptions will be further explored for robustness as outlined in Section 4.2.

As can be seen above, the population level hyper-parameter *µ*_*P*_ is shared across the phases of the clinical development. The population level model for this hyper-parameter is outlined in the next section. Since *θ*_*P* 2_, *θ*_*P* 3_ come from a Normal distribution with the same mean parameter and different levels of uncertainty, we assume that these are partially exchangeable. It is important to note that, exchangeability is a qualitative assumption and cannot be evaluated statistically (Neuenschwander et al., 2010).

### 2.2 Population Level Model

We assume that the population level hyper-parameter, *µ*_*P*_ , follows a Normal mixture distribution:

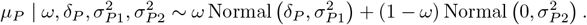

The first piece in the above mixture, Normal 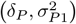, represents an enthusiastic component, with *δ*_*P*_ being the target treatment effect, i.e., the value set under the alternative hypothesis for the Phase III study power calculation. The second piece, Normal 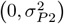 , represents a skeptical component that is centered at 0, i.e., lack of treatment effect or the value under the null. The probability that *µ*_*P*_ comes from the enthusiastic component is determined by the optimistic factor, *ω*, which is also referred to as a benchmark of PoS in this paper and is specified in Section 2.3.

Since the distributions of the enthusiastic and skeptical components are associated with the alternative and null hypothesis of the treatment effect respectively, the values representing their uncertainty, 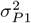 and 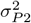 respectively, are obtained by solving the following equations:

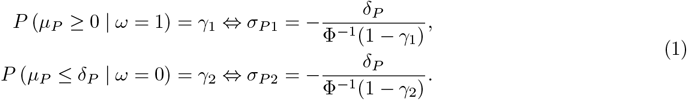

Based on (1), Figure 1 illustrates the relationship between 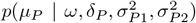 and *µ*_*P*_ for a range of values of *µ*_*P*_ , under four different settings of *ω* and *γ*_1_, *γ*_2_. The gray areas under the curve tail correspond with the tail probability defined in (1). The Figure shows that we can control the tail probabilities of the two mixture components by setting the values of *γ*_1_ and *γ*_2_ using the formula in (1).

**Figure 1.**
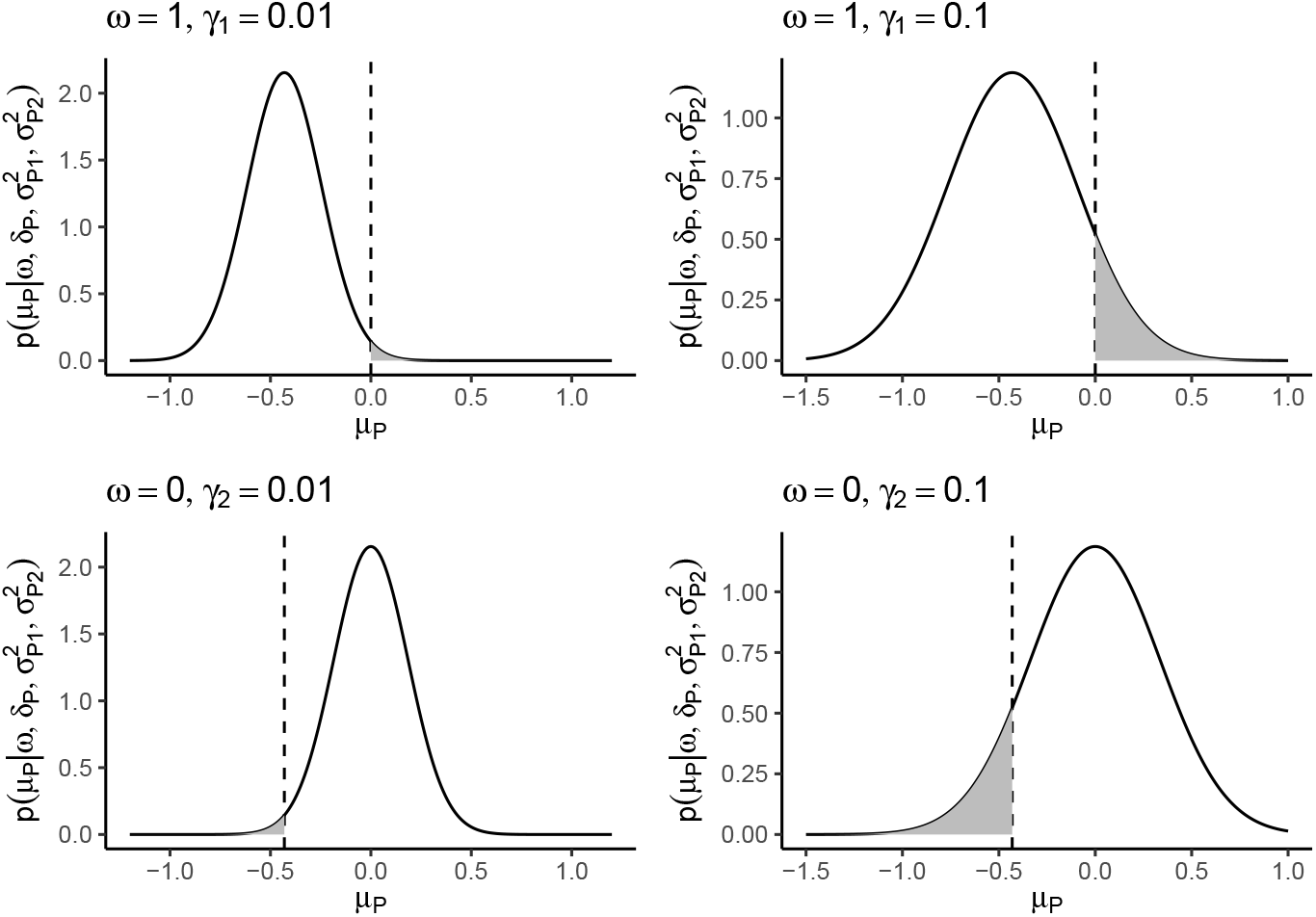
An illustration of the function relationship between 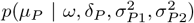, and *µ*_*P*_ under different settings of *ω* and *γ*_1_, *γ*_2_, where *δ*_*P*_ = log(0.65). The *x*-axis shows a series of equally spaced values of *µ*_*P*_ , the y-axis shows the corresponding values of 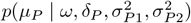. The vertical dashed line represents the threshold for the tail probability (0 for the top row and log(0.65) for the bottom row). The grey areas under the curve corresponds to the tail probabilities, i.e. *P* (*µ*_*P*_ *≤ δ*_*P*_ | *ω* = 0) and *P* (*µ*_*P*_ *≥* 0 | *ω* = 1).

### 2.3 Benchmark Estimate of PoS

Ideally the benchmark value of PoS should be estimated using historical data to obtain the probability of historical Phase III trial successes. However, the access to such data is not always easily available. Therefore, alternative approach might be needed to approximate it. For the framework presented here, the benchmark value of PoS, *ω*, was obtained from historical data by first estimating the probability of FDA approval and then converting that value to the probability of a Phase III success (e.g., meeting study objectives). Here, historical data “success” refers to whether a clinical trial and corresponding experimental therapy will eventually result in FDA regulatory approval. For example, “success” for a Phase III study means that the primary hypotheses were met followed by formal FDA approval. Let *Y* and **X** denote the outcome (FDA Approval: 1 (Yes) vs 0 (No)) and trial-level covariate space respectively. The goal is to estimate *Prob*(*Y* = 1|**X**), or the probability that a trial will lead to FDA approval conditional on the trial-level characteristics (e.g., tumor indication, phase, etc). This involves fitting some model *Y ∼ f* (**X**), where *f* (*·*) is a model that outputs probability estimates. For this paper, random forest was used to obtain probability estimates. Given the fitted model, probability estimates can be obtained for specific trial characteristics.

Let **X**_-ph_ denote the trial-level characteristics excluding the phase variable. Of particular interest is *Prob*(*Y* = 1|**X**_-ph_, *phase* = 3), or the probability of FDA approval conditional on the trial being at Phase III. Given this, *ω* can be approximated by *Prob*(*Y* = 1|**X**_-ph_, *phase* = 3)*/*0.92, where 0.92 is the average probability that a Phase III trial receives regulatory approval after meeting the study objectives (IQVIA report, 2024).

In general, the probability of FDA approval can either be used directly or adjusted by a different constant. Since the aim of generating the Phase III PoS is to enable systematic comparisons across various compounds and designs based on the available data, the precise value of *ω* is less critical; what matters more is that a consistent approach is applied to the studies being compared.

### 2.4 Phase III PoS Prediction

After fitting the models that are outlined in Sections 2.1 - 2.3, we can generate a Phase III efficacy PoS prediction based on the distribution of *θ*_*P* 3_. Since pivotal trials in oncology often consider group sequential design (GSD) with multiple analysis, we use *J* to denote the number of analyses for a future Phase III study. For instance, if *J* = 2, this means that a study has one interim analysis (IA) and one final analysis (FA). Following previously introduced notations, the distribution of the observed log HR for endpoint *P* at the *j*-th analysis, 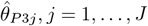, *J* is as follows:

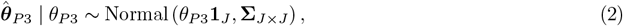

where *θ*_*P* 3_ is the underlying true log hazard ratio for all J analyses. **Σ** is the covariance matrix that encodes the Fisher’s information for 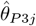 :

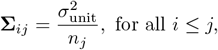

here *n*_*j*_ is the total number of events that are observed at the *j*-th analysis, and 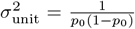, where *p*_0_ is the proportion of patients in the control group. Note that *n*_*j*_ and *p*_0_ should be set to their target values if the PoS prediction is carried out at the design stage. Specifically, *n*_*j*_ should be set to the target number of events at the *j*-th analysis and *p*_0_ should be set to the planned proportion of patients in the control group (e.g., *p*_0_ = 0.5 if the planned randomization ratio is 1 : 1). We apply the Monte Carlo simulation approach for PoS prediction. Following (2), we generate the predicted Phase III treatment effect on endpoint *P* for a total of *L* times, and let 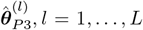, *L* be the *l*-th prediction. We let 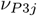 be the frequentist efficacy boundary associated with success at the *j*-th analysis (i.e., the trial is stopped for efficacy at the *j*-th analysis if the treatment effect crosses the threshold for the first time at such analysis). Thus, the probability of stopping a Phase III trial for efficacy at the first IA is estimated as

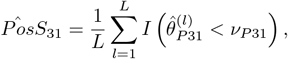

and the probability of stopping a Phase III trial for efficacy at the *j*-th analysis is estimated as

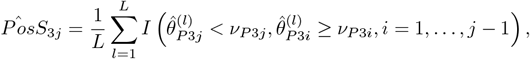

finally, the overall *PoS* is estimated as

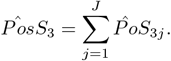

## 3 Further considerations for POS estimation

### 3.1 Study level model when Phase III primary endpoint is not available from earlier study(ies)

In this section, we consider the scenario in which a Phase III primary endpoint is not available from an earlier study(ies). This could be due to various reasons, such as a short follow-up duration in the earlier study(ies). For illustrative purposes, we assume that a Phase III primary endpoint is PFS, i.e., *P* 3 = *PFS* and *θ*_*P* 3_ = *θ*_*P F S*,3_. Also, suppose that only objective response rate (ORR) is available from the preceding randomized controlled Phase II study, i.e., *P* 2 = *ORR, θ*_*P* 2_ = *θ*_*ORR*,2_, where *θ*_*ORR*,2_ represents the log odds ratio (*OR*) of the treatment effect on ORR. We assume the Phase II observed treatment effect on ORR has a Normal distribution as follows:

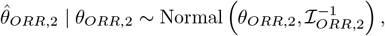

where ℐ_*ORR*,2_ is the Fisher information of 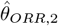. In order to infer a distribution for *θ*_*P F S*,2_ conditioning on *θ*_*ORR*,2_, we assume a linear correlation between *θ*_*ORR*,2_ and *θ*_*P F S*,2_ for simplicity. Note that such assumption is usually valid in practice based on historical data and the correlation could be estimated based on study level aggregated data meta-analysis and not patient level data.

As an example, we will use the meta-analysis on advanced non-small-cell lung cancer (NSCLC) that was reported by Blumenthal et al. (2015), and will assume the following linear model between the PFS and ORR treatment effects:

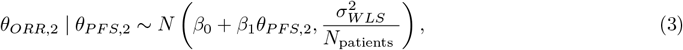

where *N*_*patients*_ is the number of patients in a given trial, and the regression parameters are assigned with the following priors:

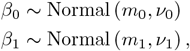

where values of the hyperparameters *m*_0_, *m*_1_, *ν*_0_, *ν*_1_ and 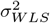 are determined by the meta-analysis regression estimates in Blumenthal et al. (2015). We defer the readers to Section 4 for details on choosing the hyperparameter values. While the distribution of *θ*_*ORR*,2_ is specific for NSCLC, a patient population that is described in the case study in Section 4, in general, different approaches could be adopted for specifying a relationship between *P* and *D*. This type of assessment is outside of the scope of this paper.

### 3.2 Study level model when an earlier study(ies) is a single arm

If an earlier study is a single arm one, then the treatment effect could be approximated based on historical data for a standard of care (SOC). We will continue to focus on ORR and PFS in this section, as these are commonly used endpoints in early oncology and serve as important references for designing pivotal trials. We will also concentrate on an aggregate study level data. However, if individual patient level data or aggregate level covariate data is available, the estimation of the treatment effect from the earlier study can incorporate differences in baseline characteristics between the single-arm study and its synthetic control through propensity methods (e.g. matching) or regression methods. This adjusted approach can yield more reliable estimates, as historical data can be adjusted such that it is more similar to the target clinical trial. Below we present an unadjusted approach. For either unadjusted or adjusted approaches, the key required output is an estimated treatment difference and corresponding standard error (SE).

#### 3.2.1 Single arm trial: ORR data is available

Let *x*_*ORR*,*trt*_ and *n*_*trt*_ be the number of responders and the total sample size in the earlier single arm trial respectively (or one of its cohorts, if applicable). Also, let *x*_*ORR*,*soc*_ and *n*_*soc*_ be the number of responders and the total sample size respectively that were previously reported in literature for an SOC, or comparator that is being considered for a future pivotal trial. The observed treatment effect and it’s SE can then be estimated approximately as:

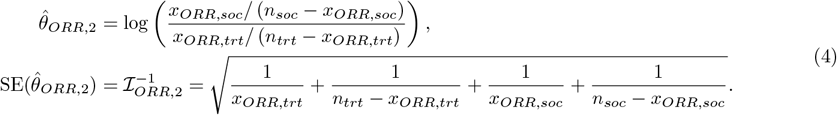

#### 3.2.2 Single arm trial: PFS is available

Let *x*_*P F S,trt*_ and *n*_*P F S,trt*_ be the median PFS and the total number of PFS events in the earlier single arm trial respectively (or one of its cohorts, if applicable). Also, let *x*_*P F S,soc*_ and *n*_*P F S,soc*_ be the median PFS and the total number of PFS events respectively that were previously reported in literature for an SOC, or comparator that is being considered for a future pivotal trial. Under proportional hazards and exponential distribution assumptions, and given that a median PFS was reached in a single arm trial and is available from the literature, the observed treatment effect and its SE can then be estimated approximately as:

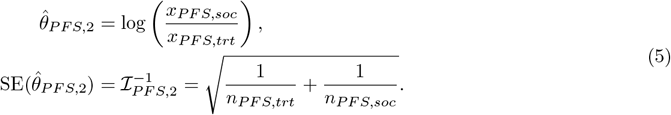

## 4 Case Study

For the case study we will use a Phase III study (PHASE3 hereafter), which was a randomized study in NSCLC that compared experimental treatment to an SOC. Both PFS and OS were primary endpoints; however, for simplicity we will focus on PFS for the PoS estimation. Table 1 shows the design assumptions for the PHASE3 protocol. Note that the target number of 468 events at the FA provides approximately 95.5% power to detect a hazard ratio of 0.7 at *α* = 0.025.

**Table 1:**
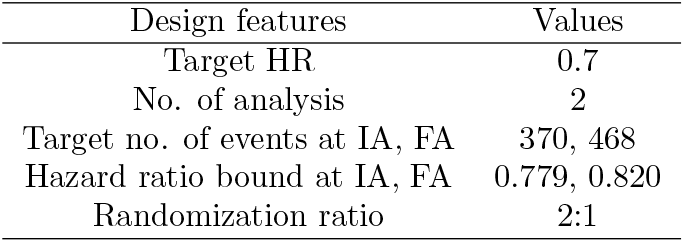
Design assumptions used in the PHASE3 protocol.

| Design features | Values |
| --- | --- |
| Target HR | 0.7 |
| No. of analysis | 2 |
| Target no. of events at IA, FA | 370, 468 |
| Hazard ratio bound at IA, FA | 0.779, 0.820 |
| Randomization ratio | 2:1 |

PHASE3 was informed by a randomized Phase II study (PHASE2 hereafter), which was conducted in a similar patient population as PHASE3 and had similar treatment groups. The PHASE2 study results are listed in Table 2. The probability of FDA approval was estimated as 0.52 based on a random forest classifier model and therefore the benchmark PoS, *ω*, was set to 0.52/0.92 = 0.57. To assess the robustness of the PoS estimates, we varied the values of the optimistic factor in the mixture prior, ranging from 0.5 to 0.65 by intervals of 0.05 in addition to the benchmark.

**Table 2:** PHASE2 study results.

| Study results | Values |
| --- | --- |
| No. of patients | 60 (Treatment)<br>63 (SOC) |
| No. of responders | 33 (Treatment)<br>18 (SOC) |
| PFS HR (95% CI) | 0.53 (0.31, 0.91) |
| No. of PFS events | 56 |

In order to further study the performance of the PoS estimates using the proposed framework, we evaluated the impact of varying model parameters values through simulations outlined in Table 3. PoS estimates will be presented for IA and FA for each simulation.

**Table 3:** Simulation studies and their corresponding section numbers.

| Sections | Original PoS estimation assumptions | Variable Factors |
| --- | --- | --- |
| Section 4.1 | Only PHASE2 PFS is used | Different PHASE2 endpoints (ORR only, both PFS and ORR) |
| Section 4.2 | ‘Very small’ PHASE3 heterogeneity | ‘Moderate’ and ‘small’ PHASE3 heterogeneity |
| Section 4.3 | PHASE2 PFS HR estimate uncertainty is reflected by 56 observed events | Increase/decrease the uncertainty by varying the number of observed PFS events to 40/70 |
| Section 4.3 | PHASE2 ORR treatment effect uncertainty is reflected by 33 responders | Increase/decrease the uncertainty by varying the number responders to 18/45 |
| Section 4.4 | ORR-PFS relationship according to (3) | Varying slope coefficients in (3) |
| Section 4.5 | Both prior and data affect PoS estimation | Prior predictive checks without data |

In scenarios where we utilize ORR data from PHASE2 for the PoS estimation, equation (3) is employed to model a positive correlation between log (HR-PFS) and (log OR-ORR). To choose the hyperparameter values in (3), (*m*_0_, *m*_1_) are point estimates for the intercept and slope from a weighted least squares (WLS) linear regression model, while (*ν*_0_, *ν*_1_) are the respective standard errors and 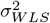 is estimated based on WLS regression residual variance.

The Bayesian hierarchical model in the proposed framework is implemented using RStan in oncoPoS package that was prepared as an integral part of this paper and is available on XXX. We run 4 Markov Chain Monte Carlo (MCMC) chains, each with 20,000 iterations, including a 50% warm-up period and 50% sampling.

### 4.1 PHASE3 PoS using different PHASE2 endpoints results

In certain situations, ORR could be the only mature endpoint in an earlier study to show anti-tumor activity when a pivotal study is being designed. In this section, we investigate how PoS estimate changes under scenarios where different PHASE2 endpoints results are included in the model. Table 4 shows the PoS estimates using three different categories of PHASE2 data: PFS results only, ORR results only, and both ORR and PFS results. The PHASE3 PoS estimates at the FA given the benchmark PoS *ω* value of 0.57 are 0.71, 0.83, and 0.87 when ORR, PFS, and both ORR and PFS results from PHASE2 are used respectively. For the same type of PHASE2 data used, PoS is generally lower for *ω* values that are below the benchmark PoS and higher for those above the benchmark PoS. In addition, the PoS is higher across different values of *ω* when both PFS and ORR results from PHASE2 are used compared to using results from only one PHASE2 endpoint. This is not surprising because both PFS and ORR results from PHASE2 indicate a substantial treatment benefit from the experimental group compared to the control.

**Table 4:** PoS of PHASE3 based on different PHASE2 endpoints results.

| PHASE2<br>Results<br>Used | PHASE3 PoS Estimate Incorporating PHASE2 Results |  |  |  |  |  |  |  |  |  |
| --- | --- | --- | --- | --- | --- | --- | --- | --- | --- | --- |
| | Below Benchmark PoS ($\omega$) | | | | Benchmark PoS ($\omega$)<sup>1</sup> | | Above Benchmark PoS ($\omega$) | | | |
|  | 0.5 |  | 0.55 |  | 0.57 |  | 0.6 |  | 0.65 |  |
|  | IA | FA | IA | FA | IA | FA | IA | FA | IA | FA |
| PFS | 0.74 | 0.81 | 0.76 | 0.83 | **0.76** | **0.83** | 0.77 | 0.84 | 0.79 | 0.86 |
| ORR | 0.58 | 0.67 | 0.60 | 0.69 | **0.62** | **0.71** | 0.64 | 0.73 | 0.65 | 0.74 |
| PFS & ORR | 0.77 | 0.85 | 0.79 | 0.87 | **0.80** | **0.87** | 0.80 | 0.87 | 0.81 | 0.88 |

<sup>1</sup> Estimated based on probability of FDA approvability that was adjusted by the average probability that a Phase III trial receives regulatory approval after meeting the study objectives.

### 4.2 PHASE3 PoS using different degree of Phase III heterogeneity

As mentioned in Section 2.1, we assume 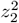 and 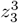 represent ‘small’ and ‘very small’ heterogeneities for treatment effects across earlier studies and Phase III studies respectively. In practice, the level of heterogeneity assumed for a Phase III study may need to be higher than for an earlier study, depending on the target study population, e.g., an earlier study focused on a specific biomarker group and Phase III allows recruitment of all-comers. To better assess the impact of this assumption, Figure 2 shows the how different degrees of Phase III heterogeneity affect the original PHASE3 POS estimate that is using PFS PHASE2 results. As expected, higher heterogeneity tends to result in lower PoS estimates at each analysis and across different benchmark estimates compared to the original PHASE3 POS. In addition, the differences are more pronounced for the FA compared to the IA for moderate heterogeneity. Similar results were observed when assessing PHASE3 POS estimates using only PFS and ORR or only ORR PHASE2 results (see Appendix A).

**Figure 2.**
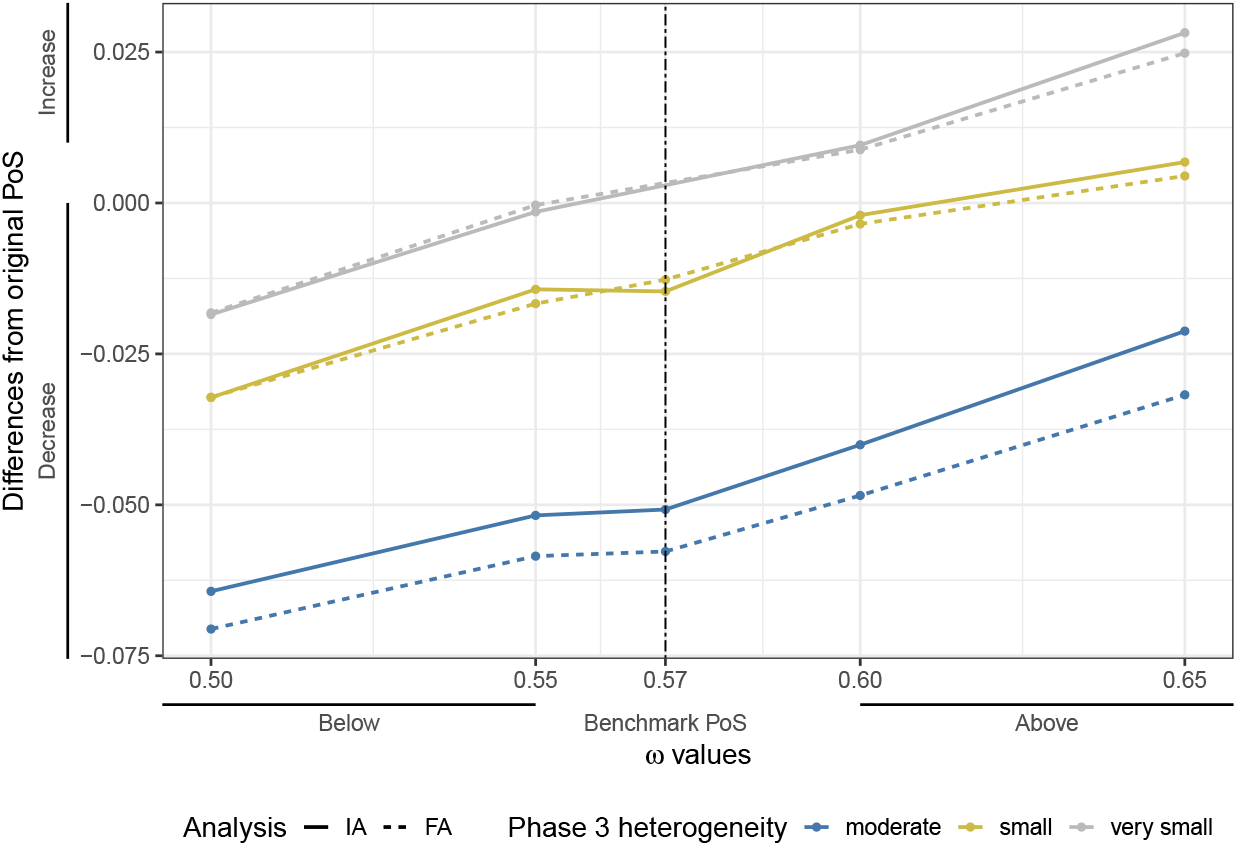
Differences from the original PHASE3 PoS using PFS PHASE2 results and based on different degrees of Phase III heterogeneity. The dashed vertical line refers to the benchmark PoS *ω* value.

### 4.3 PHASE3 PoS with varying uncertainty in PHASE2 results

As stated previously, the use of the proposed framework incorporates the uncertainty of the previously observed treatment effect. For PFS, the uncertainty of the treatment effect depends on the number of observed PFS events, while for ORR, it depends on the number of responders.To assess how such uncertainty affects PoS, we consider hypothetical scenarios for PFS and ORR PHASE2 results. In both cases the observed treatment effect for each endpoint in PHASE2 is kept the same as in the original PHASE2 study, while either the number of observed PFS events varies or number if responders in the treatment arm varies.

For PFS we examine two scenarios where the number of PFS events are 40 and 70, representing situations with more and less uncertainty in the PFS treatment effect respectively. Note that the standard error for log HR is derived using (5). As shown in Figure 3 (A), at each analysis and across different values of *ω* more PFS events, i.e., less uncertainty in the PFS treatment effect estimate, correspond to increase in the PHASE3 PoS estimates that are based on PFS PHASE2 results only.

**Figure 3.**
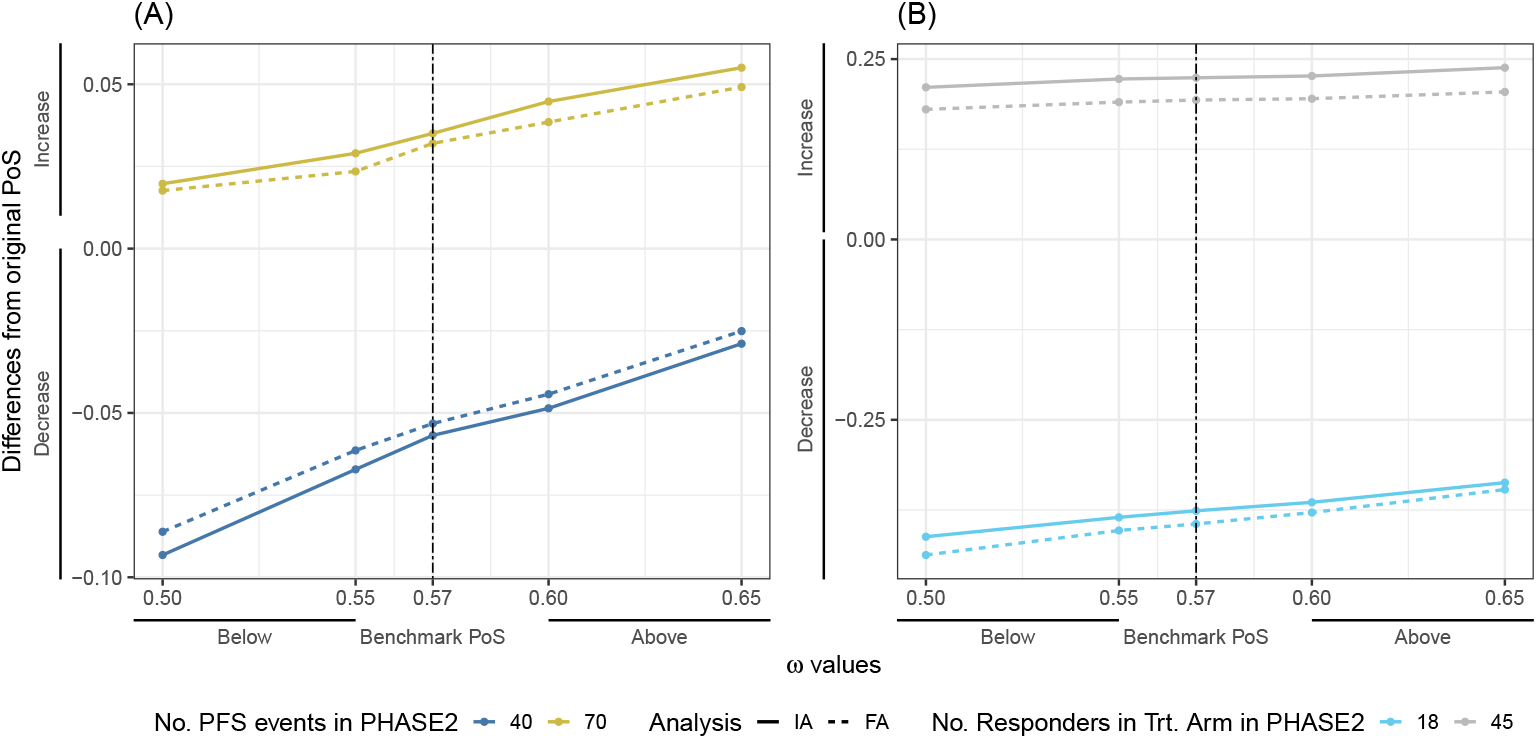
Differences from the original PHASE3 PoS (A) using PFS PHASE2 results and varying the number of PFS events (B) using ORR PHASE2 results and varying the number of responders. The dashed vertical line refers to the benchmark PoS *ω* value.

Similar to PFS, for ORR we examine two scenarios, where the number of responders in the treatment arm is assumed to be 18 and 45, representing more and less uncertainty than originally observed with the 33 responders respectively. In Figure 3 (B), we can see that at each analysis and across different values of *ω* a higher number of responders, i.e., less uncertainty in the ORR treatment effect estimate, correspond to increase in the PHASE3 PoS estimates that are based on ORR PHASE2 results only.

Also, for both above assessments, the increase in the PoS is slightly higher for the IA than for the FA for lower uncertainty, while the decrease is slightly lower for the IA than for the FA for higher uncertainty.

### 4.4 PHASE3 PoS under ORR-PFS model mis-specification

In the original PHASE3 PoS estimation, we assumed the ORR-PFS relationship follows (3), and now we evaluate how a mis-specification of the ORR-PFS model would affect the PoS estimate when only ORR results are available. Compared to the default setting, we investigate three additional hypothetical scenarios with different directions of mis-specification. To do this, we multiply the original slope parameter by factors of 0.5 (representing a case of ‘decrease’), 2 (a case of ‘increase’) and 5 (an extreme case of ‘significant increase’). These factors reflect a range of correlations between ORR and PFS, varying from stronger to weaker compared to the original relationship. The magnitude of the phase II PFS treatment effect that is derived based on the ORR data decreases as the slope parameter value in the ORR-PFS linear model increases. Therefore, it’s anticipated that a smaller slope parameter would result in a higher PoS value. Figure 4 shows the differences in the PHASE3 PoS estimates compared to the original PHASE3 POS using both PFS and ORR PHASE2 results under the 4 scenarios. We observe that the change of PoS is contigent on the direction of mis-specification. Specifically, the PoS value is inclined to be greater when the slope parameter is smaller. Moreover, the change is particularly sensitive to extreme underestimation of the phase II PFS effect when the correlation between ORR and PFS is significantly weaker than the original value (i.e., ‘significant increase’ of the slope parameter). These observations hold at both analyses and across varying values of *ω*.

**Figure 4.**
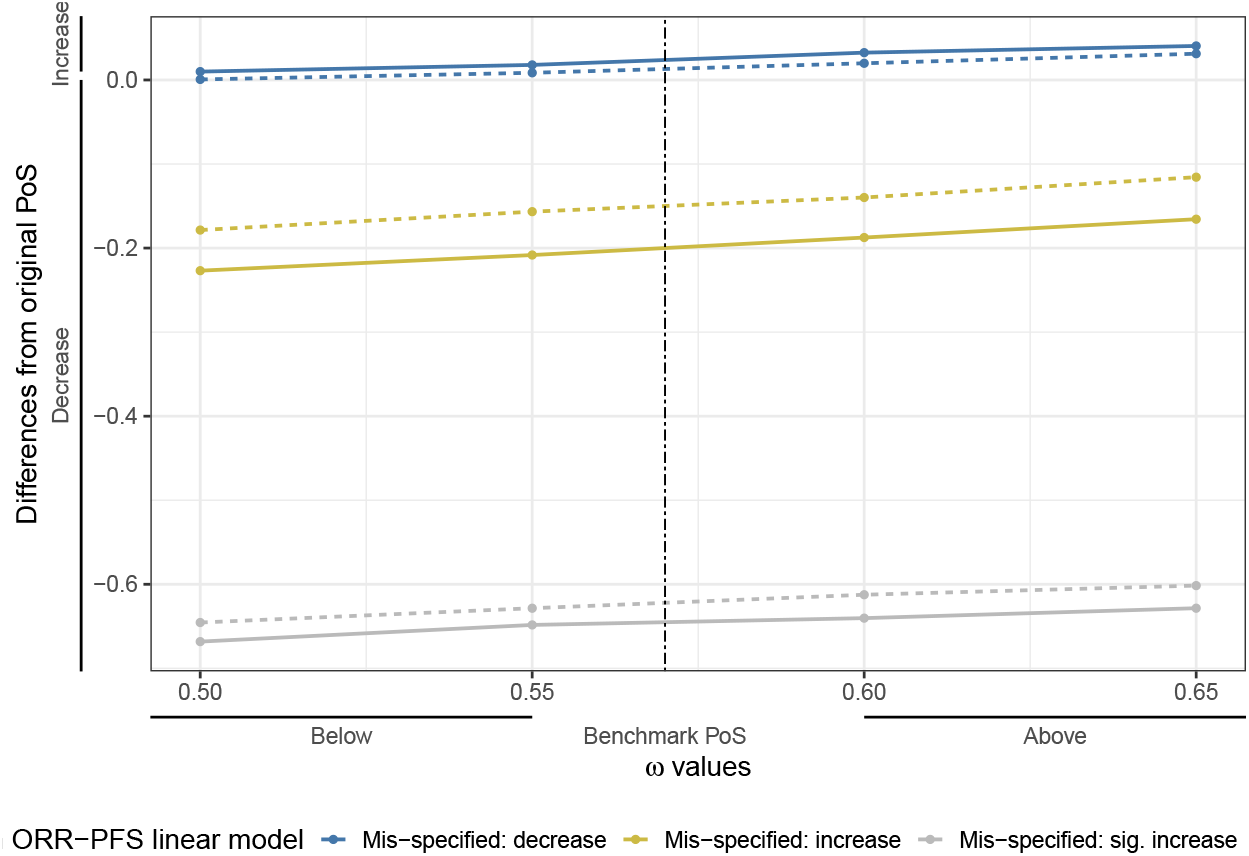
Differences from the original PHASE3 PoS using PFS and ORR PHASE2 results and based on different ORR-PFS correlation model mis-specifications. The dashed vertical line refers to the benchmark PoS *ω* value.

### 4.5 Prior predictive checks for PHASE3 when no data is used

Finally, we conduct a sanity check where no data is used, i.e., the result is only affected by the model priors. Table 5 and Table 6 shows the PoS estimates and standard errors with their associated *α*-levels and powers, when the *ω* takes values 0 and 1 respectively. We see that the PoS is close to the target alpha level at each analysis when the skeptical component is assigned a 100% weight, and close to estimated power at each analysis when the enthusisatic component is assigned a 100% weight.

**Table 5:** Prior predictive checks when the skeptical component is assigned a 100% weight (*ω* = 0)

| Analysis | Estimated PoS | SE of PoS | PHASE3 $\alpha$ Level |
| --- | --- | --- | --- |
| 1 | 0.01095 | $7.35 \times 10^{-4}$ | 0.0117 |
| 2 | 0.02525 | $1.10 \times 10^{-3}$ | 0.0250 |

**Table 6:** Prior predictive checks when the enthusiastic component is assigned a 100% weight (*ω* = 1)

| Analysis | Estimated PoS | SE of PoS | PHASE3 Power |
| --- | --- | --- | --- |
| 1 | 0.84835 | $2.53 \times 10^{-3}$ | 0.8439 |
| 2 | 0.94850 | $1.56 \times 10^{-3}$ | 0.9550 |

## 5 Discussion

In this work, we introduced a practical, coherent and transparent Bayesian decision-making framework for estimating the PoS for late-stage oncology studies. Similar to the previous work by Hampson et al. (2022), our framework integrates the uncertainty associated with the target treatment effect using Bayesian Hierarchical modeling and employs benchmark estimates to establish the population level prior for treatment effect hyper-parameter, *µ*. We have expanded upon this concept by providing additional details on the implementation of this framework in oncology trials and provided the oncoPoS R package to generate PoS numbers. The MCMC method is computationally efficient and generates the PoS numbers within 7 seconds using default settings. Our work is adaptable to scenarios when Phase III and the earlier studies use either the same or different endpoints and can be extended to accommodate the use of a single arm earlier study to estimate PoS. Unlike Hampson et al. (2022), we did not include show stoppers in our framework, considering that primary endpoints in pivotal oncology trials typically incorporate survival in the endpoints definition, which includes death due to malignancy or fatal adverse events (AEs). Furthermore, we streamlined the specification of the population level prior by directly utilizing the benchmark PoS estimate as the weight of the enthusiastic prior component, *ω*, in the distribution of *µ*.

While the exact value of *ω* is not significant, caution is essential when selecting an overly optimistic estimate, as this could mislead the PoS estimation and result in the initiation of a Phase III study that is unlikely to succeed. Therefore, *ω* may also be further refined based on feedback from multiple stakeholders. Our primary objective is to offer a systematic approach for comparing various compounds and study designs to aid decision-makers.

However, to effectively facilitate decision-making for resource allocation within a company or therapeutic area, it is crucial not to alter assumptions for the sake of generating a ‘reasonable’ PoS number. Instead, teams should adhere to consistent principles when making assumptions to ensure a fair assessment and comparison across different compounds and study designs. Preferably, an independent technical review committee can be formed outside of the study team to review the assumptions. This is important because the PoS number, on its own offers little information beyond what a power number provides. More specifically, the following four principles should be followed:

1. The predictive model for estimating benchmark PoS should be applied consistently based on relevant database of similar time period to minimize the temporal effect and differences due to model performance;
2. Similar to considerations for Phase III study power calculation, the target treatment effect for the mixture prior for population level treatment effect should be chosen carefully and consistent with the Phase III target treatment effect assumption;
3. When an early or intermediate endpoint is used in earlier studies as often seen in oncology, an appropriate prediction model should be used to translate the treatment effect from early/intermediate endpoint to the longer-term endpoint such as OS. The team has the flexibility to assume linear or non-linear prediction model, but it is essential to provide clear justifications for these assumptions to the decision makers. For example, in this paper, we assumed a linear relationship between ORR and PFS treatment effect based on meta-analysis, which may be true for most chemotherapies, ADCs or immunotherapies. However, there may not be a strong correlation between ORR and OS treatment effect for immunotherapies. Therefore, it is important to thoroughly assess the mechanism of action of the new treatment and the relevance of historical data used in assessing or assuming such relationship. In addition, the framework in this scenario could be easily extended to predict treatment effect of combination therapies based on monotherapy treatment effect using independent drug action model, incorporating uncertainties (Chen et al., 2020).
4. For earlier single-arm studies, when we try to estimate treatment effect by cross-trial comparison to SOC, even if we apply some matching or weighting method based on available covariates, uncollected confounding factors may still bias the cross-trial comparison results, such as patient or site selection, potential improvements in SOC due to temporal effect, etc. Hence, it may be advisable to calibrate the assumptions regarding the treatment effect from earlier single-arm studies to account for the additional uncertainties associated with cross-trial comparisons.

We showed the implementation of our framework through a case study as well as simulation studies in which we varied different model parameters. Group sequential design (GSD) was used due to its widespread use for pivotal oncology trials. Overall, our results align with the expectation that a higher Phase III PoS is associated with a stronger treatment benefit as well as lower uncertainty from the earlier studies. In addition, we showed that population heterogeneity for a Phase III treatment effect should be taken into account when evaluating the Phase III PoS. We also demonstrated that when assigning extreme weight of 0 or 1 respectively to the enthusiastic prior component, the derived PoS estimate from the proposed framework is close to alpha level and power from the original Frequentist computations, respectively.

In cases where different endpoints are used in Phase III and the earlier studies, we adapted the meta-analysis on advanced NSCLC from (Blumenthal et al., 2015) to account for the ORR-PFS relationship. The adaption was deemed appropriate for the simulation study conducted in this paper, given that our case study was based on PHASE2 and PHASE3 under the same disease indication of NSCLC, with a similar patient population. We further studied the impact of model mis-specification on PoS estimation in the context of the ORR-PFS relationship. The results suggested that the alteration of PoS is subject to the direction of mis-specification. For example, if the correlation between ORR and PFS is assumed to be weaker than it actually is, then the PoS estimate will be more conservative.

Nevertheless, in scenarios where the correlation between ORR and PFS is not established or expected for a particular therapy or indication, it becomes imperative to obtain PFS data from earlier studies to ensure that the decision-making process is grounded in robust evidence. If such data are not available, one may consider making assumptions based on the mechanism of action (MoA) or pathological similarity of current compound or indication of interest to those with established correlation, with clear communication to decision-makers on the cautions to be taken. While the meta-analyses for other patient populations are beyond the scope of this paper, they carry significant implications for the practical application of the framework discussed here. Without accurately estimating the relationship between an earlier study’s endpoint and the primary endpoint in the forthcoming Phase III study, the usefulness of prior information during the Phase III design phase is limited. Therefore, publishing more meta-analyses or collecting relevant data in diverse oncology patient populations would be highly beneficial for enhancing the robustness of these estimations.

Following Hampson et al. (2022), we assigned a half-normal prior distribution to 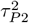 and 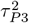 in Section 2.1. Alternatively, we can approximate the half-normal priors with inverse-Gamma priors through moment-matching equations. Using such inverse Gamma priors leads to a conjugate model that allows for Gibbs-type closed form updates with potentially more efficient posterior sampling (Levin and Peres, 2017).

Another important aspect of the framework we presented is the value of *ω*, which, as we showed, has direct implications on the on the PoS estimate. To determine this value, access to a database with relevant data is necessary for estimating the benchmark. One potential future extension of our work involves addressing the uncertainty associated with the estimated value of *ω*.

A limitation of the proposed framework is its lack of consideration for other critical factors, including regulatory and market success. Nevertheless, we anticipate that these considerations could be incorporated into the PoS assessment in future research efforts by integrating this framework with other decision making frameworks, such as analytic hierarchy process proposed by Gould et al. (2016) or multicriteria decision analysis proposed by Saint-Hilary et al. (2018) . In addition, we plan to extending this framework by using multiple endpoints for a phase III success as well as by adding futility analysis.

Considering the substantial expenses associated with phase III oncology studies, we believe that our framework is of utmost importance for resource prioritization. Wu and Ono (2021) reported that out of the 196 completed phase III randomized controlled oncology studies they extracted from ClinicalTrials.gov only 40.3% achieved the pre-specified statistical goal of the study, meaning they met the primary endpoint. As per Jardim et al. (2017), who compared 43 failed oncology development programs to 37 successful programs, the primary factors contributing to failure were the absence of biomarker-driven strategy and the lack of attaining proof of concept in earlier studies. This highlights the critical need to utilize available data and take into consideration data uncertainty to better quantify the probability of statistical success in phase III oncology studies. We hope that our framework will contribute to improving such success rate by assisting decision making for resource allocation.

## Data Availability

No new data was generated.

## 6 Acknowledgements

The authors thank Keaven Anderson, Christine Gause, and Yue Shentu for their review and comments on the statistical analysis plan and the manuscript.

## A Additional Figures

**Figure 5.**
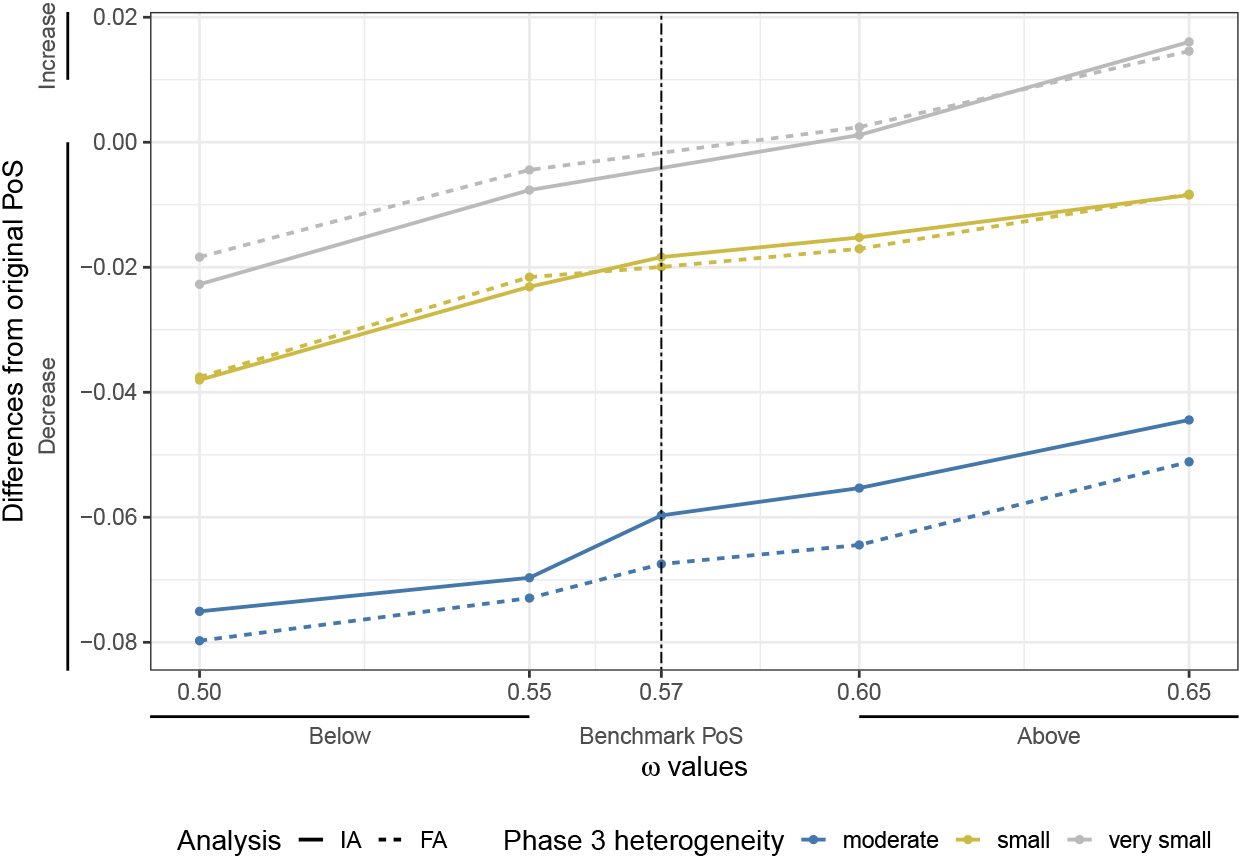
Differences from the original PHASE3 PoS using PFS & ORR PHASE2 results and based on different degrees of Phase III heterogeneity. The dashed vertical line refers to the benchmark PoS *ω* value.

**Figure 6.**
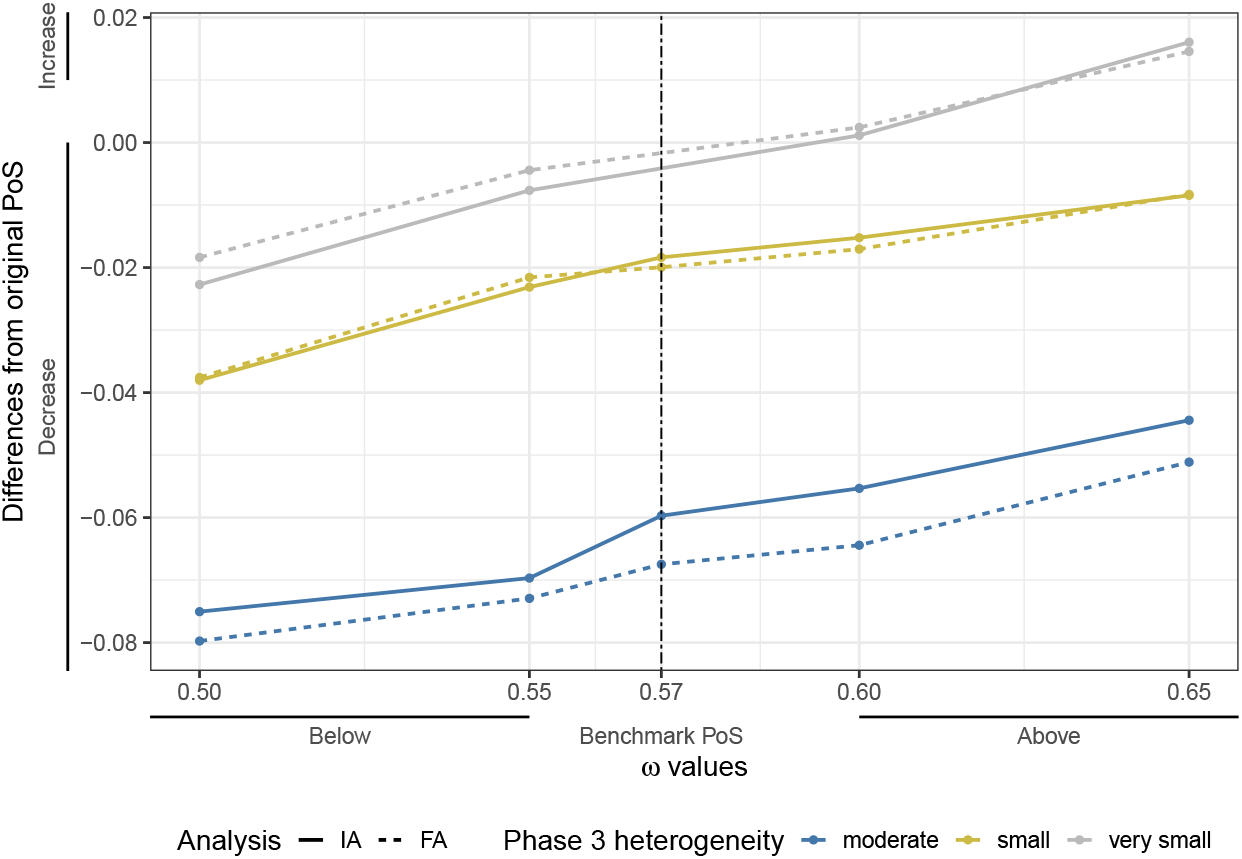
Differences from the original PHASE3 PoS using ORR PHASE2 results and based on different degrees of Phase III heterogeneity. The dashed vertical line refers to the benchmark PoS *ω* value.

## References

Blumenthal, G. M., Karuri, S. W., Zhang, H., Zhang, L., Khozin, S., Kazandjian, D., Tang, S., Sridhara, R., Keegan, P., and Pazdur, R. (2015). Overall response rate, progression-free survival, and overall survival with targeted and standard therapies in advanced non–small-cell lung cancer: Us food and drug administration trial-level and patient-level analyses. Journal of Clinical Oncology, 33(9):1008.

Chen, C., Liu, F., Ren, Y., Suttner, L., Sun, Z., Shentu, Y., and Schmidt, E. V. (2020). Independent drug action and its statistical implications for development of combination therapies. Contemporary Clinical Trials, 98:106126.

Chen, C., Zhou, X., Lavezzi, S. M., Arshad, U., and Sharma, R. (2023). Concept and application of the probability of pharmacological success (pops) as a decision tool in drug development: a position paper. Journal of Translational Medicine, 21(1):17.

Food and Drug Administration (FDA) (2021). Development & Approval Process (Drugs). https://www.fda.gov/drugs/development-approval-process-drugs. [Online; accessed October-2024].

Fougeray, R., Vidot, L., Ratta, M., Teng, Z., Skanji, D., and Saint-Hilary, G. (2024). Futility Interim Analysis Based on Probability of Success Using a Surrogate Endpoint. Pharmaceutical Statistics.

Gould, A. L., Krishna, R., Khan, A., and Saltzman, J. (2016). Incorporating clinical knowledge and experience in the evaluation of drug development projects using the analytic hierarchy process. International Journal of Business and Systems Research, 10(2-4):105–123.

Hampson, L. V., Holzhauer, B., Bornkamp, B., Kahn, J., Lange, M. R., Luo, W.-L., Singh, P., Ballerstedt, S., and Cioppa, G. D. (2022). A new comprehensive approach to assess the probability of success of development programs before pivotal trials. Clinical Pharmacology & Therapeutics, 111(5):1050–1060.

Hay, M., Thomas, D. W., Craighead, J. L., Economides, C., and Rosenthal, J. (2014). Clinical development success rates for investigational drugs. Nature biotechnology, 32(1):40–51.

Jardim, D. L., Groves, E. S., Breitfeld, P. P., and Kurzrock, R. (2017). Factors associated with failure of oncology drugs in late-stage clinical development: A systematic review. Cancer Treatment Reviews, 52:12–21.

Levin, D. A. and Peres, Y. (2017). Markov chains and mixing times, volume 107. American Mathematical Soc.

Neuenschwander, B., Capkun-Niggli, G., Branson, M., and Spiegelhalter, D. J. (2010). Summarizing historical information on controls in clinical trials. Clinical Trials, 7(1):5–18.

Proper, J. L., Bunn, V., Hupf, B., and Lin, J. (2024). Predicting Probability of Success for Phase III Trials via Propensity-Score-Based External Data Borrowing. Statistics in Biopharmaceutical Research, pages 1–13.

Saint-Hilary, G., Robert, V., and Gasparini, M. (2018). Decision-making in drug development using a composite definition of success. Pharmaceutical Statistics, 17(5):555–569.

Schlander, M., Hernandez-Villafuerte, K., Cheng, C.-Y., Mestre-Ferrandiz, J., and Baumann, M. (2021). How much does it cost to research and develop a new drug? a systematic review and assessment. Pharmacoeconomics, 39:1243–1269.

Schuhmacher, A., Hinder, M., Brief, E., Gassmann, O., and Hartl, D. (2025). Benchmarking r&d success rates of leading pharmaceutical companies: an empirical analysis of fda approvals (2006–2022). Drug Discovery Today, page 104291.

IQVIA report (2024). Global trends in R&D. https://www.iqvia.com/insights/the-iqvia-institute/reports-and-publications/reports/global-trends-in-r-and-d-2024-activity-productivity-and-enablers. [Online; accessed 10-June-2024].

Wang, Y., Fu, H., Kulkarni, P., and Kaiser, C. (2013). Evaluating and utilizing probability of study success in clinical development. Clinical Trials, 10(3):407–413.

Wong, C. H., Siah, K. W., and Lo, A. W. (2019). Estimation of clinical trial success rates and related parameters. Biostatistics, 20(2):273–286.

Wu, C. and Ono, S. (2021). Exploratory analysis of the factors associated with success rates of confirmatory randomized controlled trials in cancer drug development. Clinical and Translational Science, 14(1):260–267.

